# State-independent functional connectomic architecture captures transdiagnostic network alterations and clinical variation

**DOI:** 10.64898/2026.10.30.26366891

**Authors:** Enhui Zhang, Ruoyun Zhang, Anfu Deng, Yanlong Hou, Bihua Xu, Shaolei Guo, Jiaxi Liao, Yulin Wang, Debo Dong

**Affiliations:** Key Laboratory of Cognition and Personality, Ministry of Education, Faculty of Psychology, Southwest University, Chongqing, China; The Second Mental Health Center of Beibei District, 400715, Chongqing; Institute of Neuroscience and Medicine, Brain and Behaviour (INM-7), Research Centre Jülich, Jülich, Germany

**Keywords:** transdiagnostic psychiatry, cross-paradigm connectivity, state-independent functional architecture, functional connectome, cerebellum, clinical heterogeneity

## Abstract

**Background:** Psychiatric disorders show substantial overlap across diagnostic categories while differing markedly in clinical expression. Whether these shared abnormalities and individual differences are embedded in stable functional brain architecture rather than in state-specific connectivity remains unclear.

**Methods:** We integrated functional connectomes across one resting-state and six task-based fMRI conditions in 377 participants from the YaleNeuroConnect cohort, including 223 healthy controls and 154 individuals with psychiatric disorders. Principal component analysis was applied to each participant’ s functional connectivity data across the seven states to derive a state-independent connectivity representation. Network-based statistics then identified components showing significant case-control differences. Cross-state consistency was assessed, and findings were validated in an independent UCLA cohort of 177 participants. Associations between component expression and clinical measures were further examined within the Yale psychiatric group.

**Results:** Cross-paradigm connectivity revealed two reproducible patterns of state-independent network alteration. One pattern showed reduced connectivity among limbic, somatomotor and salience systems, whereas the other showed stronger cerebellar coupling with limbic and somatomotor systems. Both patterns showed consistent case-control differences across functional states and generalized to an independent UCLA cohort. Their clinical relevance, however, differed. Greater expression of the cerebello-cortical connectivity pattern was associated with lower positive affect, greater psychopathology and more everyday executive difficulties, whereas the reduced limbic-somatomotor-salience connectivity pattern showed no significant clinical associations.

**Conclusions:** These findings delineate a stable reorganization of large-scale functional architecture across psychiatric disorders and suggest that broadly expressed psychiatric abnormalities and individual clinical variation may be differentially represented within the altered connectome.

## Introduction

Psychiatric disorders show substantial overlap across diagnostic categories while also varying markedly in their clinical expression, posing a challenge for understanding their underlying neurobiology. Symptoms, cognitive difficulties and functional impairments frequently extend across conventional diagnostic boundaries, while individuals with the same diagnosis can differ considerably in symptom severity and everyday functioning (1,2). In line with this clinical heterogeneity, psychiatric neuroscience has increasingly moved beyond disorder-specific abnormalities toward transdiagnostic and dimensional approaches that seek neural features shared across disorders as well as neural variation related to individual differences in psychopathology (2–5). These lines of work suggest that psychiatric disorders may be better characterized by distributed abnormalities in large-scale brain systems than by isolated changes confined to individual diagnoses.

Functional connectivity has provided an important systems-level framework for this effort. Altered interactions among large-scale networks involved in salience processing, affective regulation, sensorimotor function and higher-order cognition have been reported across schizophrenia, bipolar disorde and other psychiatric conditions (6–9). Resting-state connectome studies have further identified transdiagnostic patterns that generalize across diagnostic groups and relate to dimensional psychiatric symptoms (2,10). These findings together indicate that functional connectivity captures both common psychiatric dysfunction and individual clinical variation.

Despite this progress, existing studies have been limited in several respects. First, most transdiagnostic connectome studies have examined psychiatric abnormalities within a single functional state. Functional connectivity, however, varies across cognitive contexts and reflects both relatively stable features of individual brain organization and state-dependent modulation (11,12). It therefore remains unclear whether reported transdiagnostic abnormalities represent persistent features of functional architecture or are partly specific to the state in which they were measured. Second, studies identifying shared case-control abnormalities and studies examining dimensional brain-clinical relationships have often pursued these aims separately. Whether the neural features that broadly distinguish psychiatric populations from healthy individuals are also those that account for variation in clinical expression remains insufficiently understood. Third, although cross-paradigm connectivity has been developed to isolate functional organization that is shared across cognitive states, its psychiatric applications have so far focused largely on schizophrenia and psychosis-spectrum phenotypes (13–15). Whether state-independent functional architecture reveals reproducible abnormalities across a more diagnostically heterogeneous psychiatric population remains unknown.

Cross-paradigm connectivity provides an opportunity to address these limitations by integrating connectivity acquired under multiple functional conditions and attenuating paradigm-specific variation while retaining the connectivity structure that is consistently expressed across contexts (12). Previous work has shown that such state-independent connectivity patterns can capture clinically relevant features of psychosis. Cerebello-thalamo-cortical hyperconnectivity across paradigms has been associated with psychosis vulnerability and subsequent conversion (15), while other cross-paradigm signatures have been linked to schizophrenia polygenic risk and individualized antipsychotic response (13,14). These findings highlight the utility of state-independent connectivity for stable psychiatric brain features. Applying this approach transdiagnostically may reveal which abnormalities persist across states and relate to clinical heterogeneity.

In this study, we addressed these questions using the YaleNeuroConnect dataset, which combines clinical phenotyping with resting state and task conditions. We derived participant-specific state-independent connectivity representations from one resting-state and six task-based fMRI condition. Specifically, we asked three questions. First, can state-independent connectivity identify reproducible network alterations that differentiate a diagnostically heterogeneous psychiatric population from healthy controls? Second, are these alterations consistently expressed across distinct functional states and generalizable to an independent transdiagnostic cohort with a different diagnostic composition and task battery? Third, how do individual differences in these state-independent connectivity alterations relate to clinical and cognitive functions? By addressing these questions within a single framework, we sought to characterize the stable functional architecture of psychiatric disorders and its relationship to individual clinical variation. See Fig 1 for study overview and analytical workflow.

**Fig 1.**
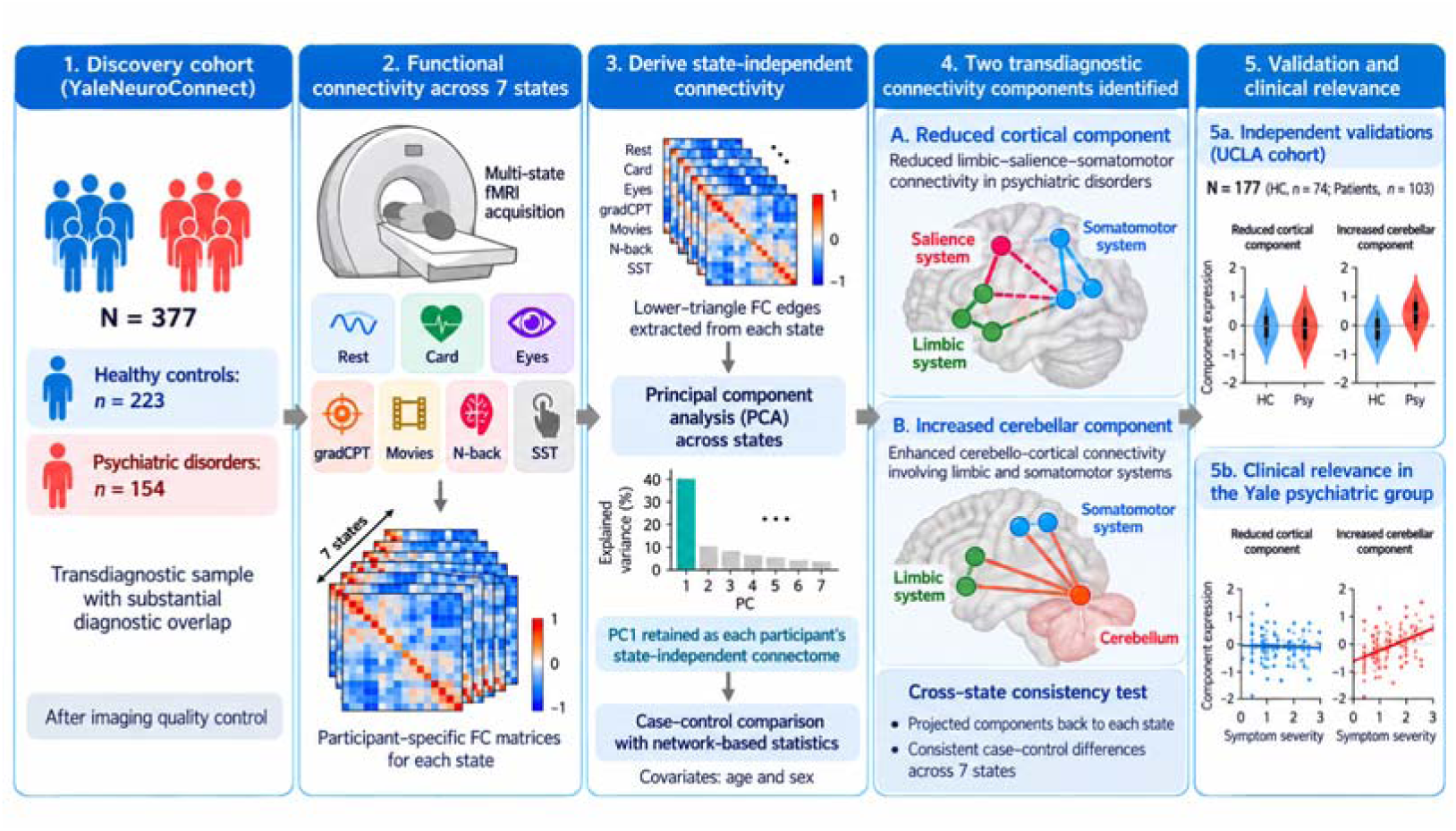
Overview of the study design and analytical workflow. Functional connectivty was examined across seven functional states in the YaleNeuroConnect discovery cohort (n = 377; 223 healthy controls and 154 individuals with psychiatric disorders). Participant-specific connectivity matrices were integrated using principal component analysis (PCA), with the first principal component (PC1) retained as the state-independent connectivity representation. Network-based statistics identified two components showing reduced limbic–salience–somatomotor connectivity and increased cerebello-cortical connectivity, respectively. Cross-state consistency was assessed by projecting the identified components onto the original connectivity matrices across all seven functional states. The two components were subsequently validated in an independent UCLA cohort (n = 177; 74 healthy controls and 103 individuals with psychiatric disorders). Finally, associations between component expression and clinical measures were examined within the Yale psychiatric group, revealing significant clinical associations for the increased cerebellar component but not the reduced cortical component.

## Methods

### Participants

We analyzed the publicly available YaleNeuroConnect dataset (16), which provides multimodal imaging and extensive clinical phenotyping in participants recruited from the local community and from Yale clinical programs. All participants gave written informed consent under a protocol approved by the Yale Institutional Review Board (HIC #2000020891).

The present analysis used the functional connectivity data released with the YaleNeuroConnect resource. Of the 410 participants in the original cohort, 33 were excluded for imaging artifacts, excessive head motion (>30% of frames with framewise displacement >0.5 mm), incomplete fMRI runs or incomplete demographic data. The final discovery sample comprised 377 participants: 223 healthy controls (112 females, 111 males; age 30.63 +/- 11.06 years) and 154 individuals with psychiatric disorders (86 females, 68 males; age 31.70 +/- 11.25 years), who did not differ in age (t = −0.91, P = 0.362) or sex distribution (chi-square = 0.94, P = 0.332). Diagnostic overlap was substantial, with 121 of 154 individuals (78.6%) meeting criteria for two or more diagnostic categories. Demographic characteristics are summarized in Table 1 and diagnostic frequencies in Table S1.

**Table 1.**
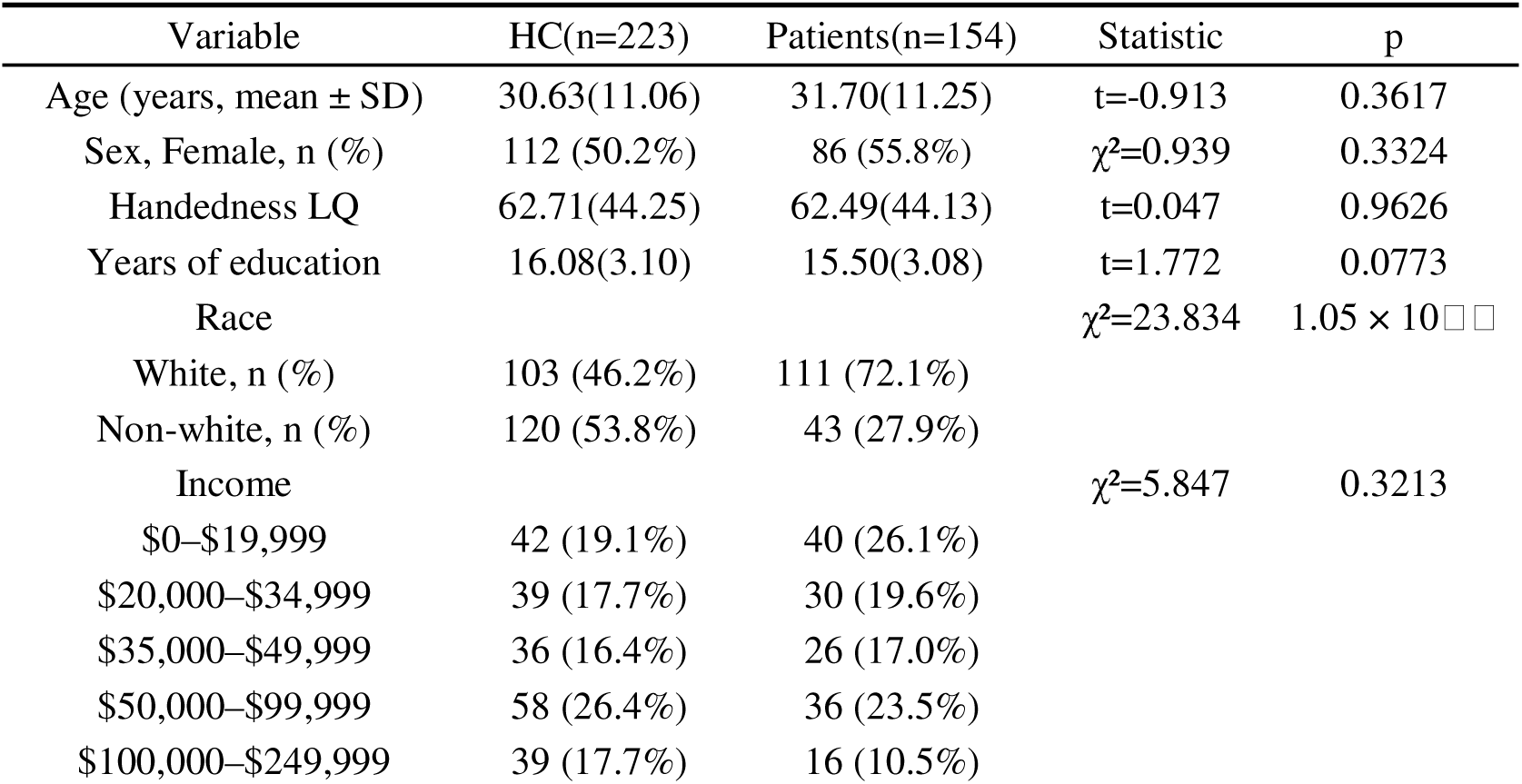
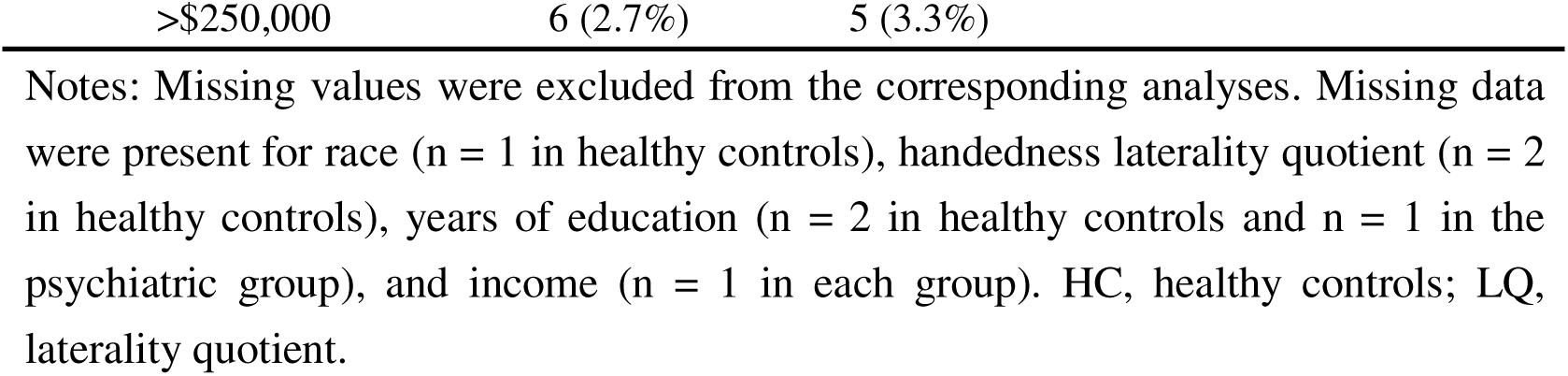
Demographic characteristics of the YaleNeuroConnect discovery cohort.

| Variable | HC(n=223) | Patients(n=154) | Statistic | p |
|---|---|---|---|---|
| Age (years, mean ± SD) | 30.63(11.06) | 31.70(11.25) | t=-0.913 | 0.3617 |
| Sex, Female, n (%) | 112 (50.2%) | 86 (55.8%) | χ²=0.939 | 0.3324 |
| Handedness LQ | 62.71(44.25) | 62.49(44.13) | t=0.047 | 0.9626 |
| Years of education | 16.08(3.10) | 15.50(3.08) | t=1.772 | 0.0773 |
| Race |  |  | χ²=23.834 | 1.05 × 10□□ |
| White, n (%) | 103 (46.2%) | 111 (72.1%) |  |  |
| Non-white, n (%) | 120 (53.8%) | 43 (27.9%) |  |  |
| Income |  |  | χ²=5.847 | 0.3213 |
| \$0–\$19,999 | 42 (19.1%) | 40 (26.1%) |  |  |
| \$20,000–\$34,999 | 39 (17.7%) | 30 (19.6%) |  |  |
| \$35,000–\$49,999 | 36 (16.4%) | 26 (17.0%) |  |  |
| \$50,000–\$99,999 | 58 (26.4%) | 36 (23.5%) |  |  |
| \$100,000–\$249,999 | 39 (17.7%) | 16 (10.5%) |  |  |
| >\$250,000 | 6 (2.7%) | 5 (3.3%) |  |  |

Notes: Missing values were excluded from the corresponding analyses. Missing data were present for race (n = 1 in healthy controls), handedness laterality quotient (n = 2 in healthy controls), years of education (n = 2 in healthy controls and n = 1 in the psychiatric group), and income (n = 1 in each group). HC, healthy controls; LQ, laterality quotient.

### Clinical and cognitive assessments

Clinical and cognitive functioning was assessed using a comprehensive battery of self-report questionnaires and standardized neuropsychological tests. Clinical measures included the Adult Temperament Questionnaire (ATQ), Behavior Rating Inventory of Executive Function–Adult Version (BRIEF-A), Brief Symptom Inventory (BSI), Interpersonal Reactivity Index (IRI), Positive and Negative Affect Schedule–Expanded Form (PANAS-X), Pittsburgh Sleep Quality Index (PSQI), and Perceived Stress Scale (PSS). Cognitive assessments included the Boston Naming Test (BNT), Wide Range Assessment of Memory and Learning (WRAML), Wechsler Adult Intelligence Scale (WAIS), Delis–Kaplan Executive Function System (D-KEFS), and Wechsler Abbreviated Scale of Intelligence (WASI). Overall, individuals with psychiatric disorders exhibited greater symptom burden and poorer cognitive performance than healthy controls (Table S2).

### MRI acquisition

See Supplementary materials for scanning parameters (17). Each participant completed two resting-state and six task runs of 6 min 49 s each, providing more than 48 min of functional imaging within an approximately 90-min session.

### fMRI conditions

During rest, participants kept their eyes open and viewed a fixation cross. Only the first resting-state run, acquired before the task battery, was analyzed, so that resting data were not influenced by preceding tasks. The six task runs comprised Card Guessing (18–20), Reading the Mind in the Eyes (21), the gradual-onset continuous performance task (22), movie watching, n-back working memory (23–25) and the stop-signal task (26,27). Task order was counterbalanced across participants and stimuli were presented using Psychtoolbox.

Together, the seven states sampled reward processing, social cognition, sustained attention, naturalistic perception, working memory, inhibitory control and unconstrained task-free activity; detailed task procedures are described in the YaleNeuroConnect data descriptor (16).

### MRI preprocessing and functional connectivity

We used the preprocessed connectivity matrices released by the YaleNeuroConnect investigators. The pipeline included skull stripping, motion correction, nonlinear normalization to MNI space, functional-to-structural registration, nuisance regression, temporal filtering and correction for head-motion effects (see Supplementary materials); scripts are available in the YaleNeuroConnect processing repository.

Functional connectivity was defined using the Shen 268-node functional atlas (28). Regional mean time series were extracted for each participant and condition, and pairwise Pearson correlations were Fisher r-to-z transformed, yielding a 268 x 268 matrix for each run. As described above, only the first resting-state run and the six task runs were analyzed.

### Cross-paradigm estimation of state-independent functional connectivity

State-independent functional connectivity was estimated using the cross-paradigm connectivity framework described by Cao (12). For each participant, the seven Fisher z-transformed matrices were vectorized across 35,778 unique edges and entered into a participant-level principal component analysis across functional states. The first principal component (PC1) was retained as the dominant pattern shared across the seven conditions and reshaped into a symmetric edgewise connectivity matrix.

We also retained the variance explained by PC1 and the contribution of each state to PC1, to verify that the dominant cross-paradigm representation was expressed similarly in both groups and was not driven by a single paradigm. The resulting participant-specific PC1 matrix is referred to as the state-independent connectivity representation.

### Case-control alterations in state-independent connectivity

To identify alterations associated with psychiatric status, we compared participant-specific state-independent matrices between groups using edgewise general linear models with age, sex and race as covariates. This analysis identifies distributed patterns that differentiate the diagnostically heterogeneous psychiatric sample from controls; diagnosis-level analyses would be required to establish how uniformly each pattern is shared across individual disorders.

Network-based statistics (NBS) controlled family-wise error over connected sets of altered edges (29). Edges surviving a primary threshold of P < 0.001 (uncorrected) were grouped into connected components, which were then evaluated using 10,000 permutations of group labels and the maximal component-size distribution (P_FWE_ < 0.05). Increased and decreased components were assessed separately, and significant edges were assigned to Shen large-scale networks for descriptive summaries.

To assess whether NBS effects were present in the original connectivity space rather than specific to the PCA representation, each significant edge mask was projected back onto the seven Fisher z-transformed matrices. Mean connectivity across the edges of each mask was computed for each participant and state, and group differences were assessed separately for each state with false-discovery-rate correction across the seven paradigms.

### Independent validation of the identified connectivity components in the UCLA CNP cohort

Generalizability was evaluated in the UCLA Consortium for Neuropsychiatric Phenotyping cohort (CNP) (30), a public dataset (OpenNeuro ds000030) that includes multiple psychiatric diagnoses and functional imaging across rest and several task conditions. Recruitment, phenotyping and consent procedures are detailed in the original publication.

After quality control, the validation sample comprised 177 participants (Table S3): 74 healthy controls and 103 individuals with psychiatric disorders (33 with schizophrenia, 38 with bipolar disorder and 32 with attention-deficit/hyperactivity disorder). Data were preprocessed with fMRIPrep 24.1.1 using procedures comparable to those applied in the discovery cohort (see Supplementary materials). Seven states were analyzed: resting state, Balloon Analogue Risk Task, paired-associate memory encoding and retrieval, stop-signal task, spatial capacity working memory and task switching (26,31–34).

The same cross-paradigm PCA was applied independently in the UCLA cohort. The two Yale components were carried forward as fixed edge masks, and mean state-independent connectivity within each mask was calculated for each participant. Case-control differences in these component-expression scores were tested with age, sex and education as covariates, and the masks were also projected onto each UCLA functional state to evaluate the consistency of effect direction.

### Clinical relevance of the identified connectivity components

For each significant NBS component, a component-expression score was calculated as the mean state-independent connectivity across its edges, yielding one score per component for each participant. Within the psychiatric group of the discovery cohort, partial correlations related these scores to the available clinical and cognitive measures, controlling for age, sex and race; two-sided P values were false-discovery-rate corrected across the tested associations.

## Results

### State-independent functional architecture across seven functional states

Across the seven analyzed functional states, PC1 accounted for 56.16% of the total variance in the discovery cohort. This dominant component therefore captured more than half of the cross-state variation in functional connectivity and was retained as the participant-level state-independent connectivity representation.

The proportion of variance explained by PC1 did not differ between the psychiatric and healthy-control groups (t = 1.084, P = 0.279). Contributions from the individual paradigms were also comparable between groups; no paradigm-specific PC1 loading survived correction for multiple comparisons (all FDR-adjusted P > 0.35). These results indicate that subsequent group differences were not attributable to a gross difference in the overall contribution of one functional state to the cross-paradigm representation.

### State-independent connectivity alterations in the transdiagnostic discovery cohort

NBS identified two distributed connectivity components that differed between the psychiatric and healthy-control groups. One component showed lower state-independent connectivity in the psychiatric group, whereas the other showed higher connectivity. The two patterns involved different large-scale network configurations rather than a uniform whole-brain shift in connectivity.

The decreased-connectivity component comprised 66 edges connecting 35 nodes (P_FWE_ < 0.05), with alterations predominantly involving limbic, somatomotor and salience systems (Fig. 2a). The most prominent connections linked somatomotor and limbic regions, with additional involvement of limbic-salience and somatomotor-salience coupling.

**Fig. 2.**
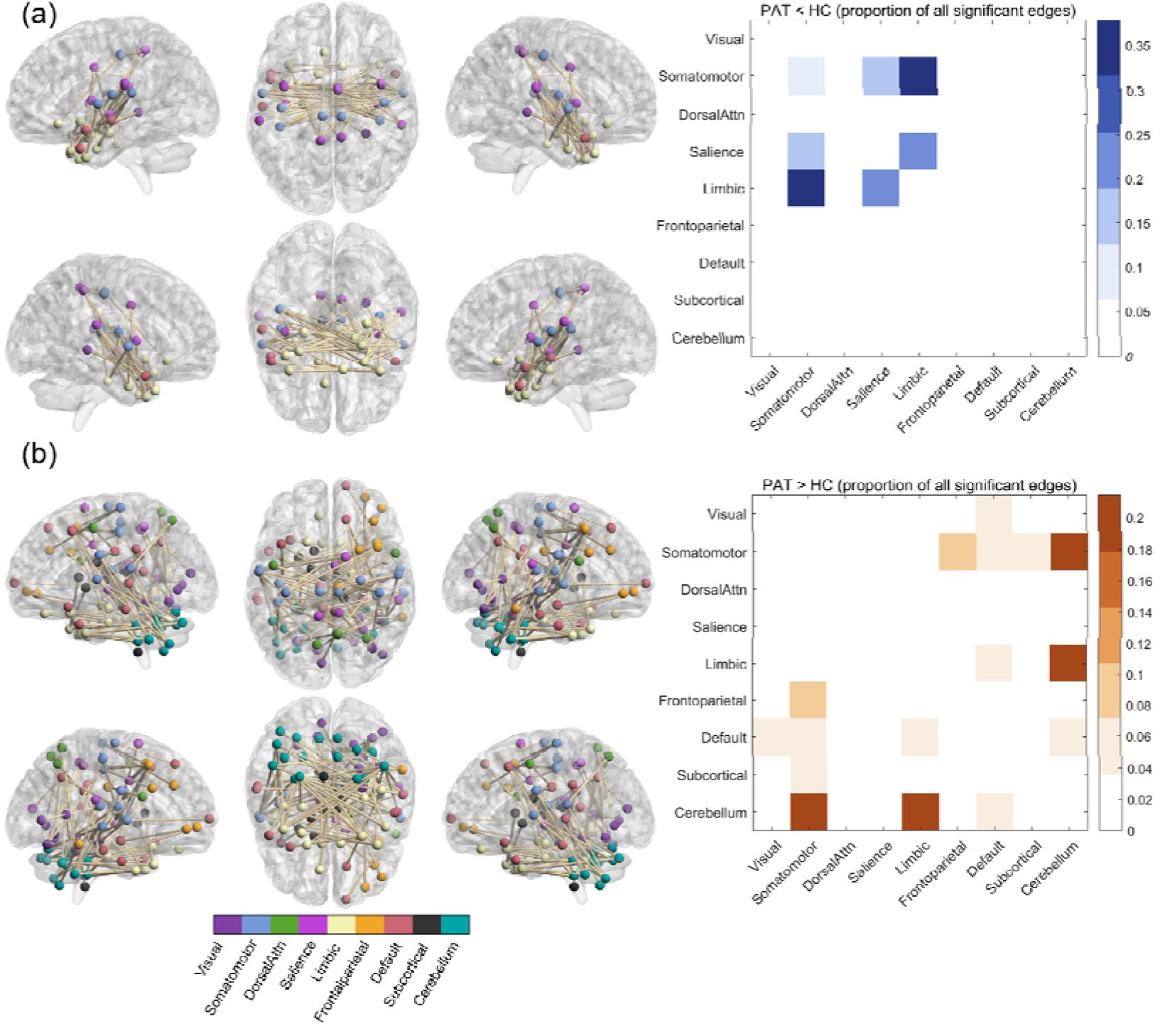
State-independent functional connectivity alterations in the psychiatric group. **a**, Network-based statistics identified a connected component showing lower state-independent functional connectivity in participants with psychiatric disorders than in healthy controls. The spatial distribution of significant edges and their network assignments is shown together with the relative distribution of significant edges across large-scale network pairs. Reduced connectivity was concentrated among limbic, somatomotor, and salience systems. **b**, A second connected component showed higher state-independent functional connectivity in participants with psychiatric disorders than in healthy controls. Increased connectivity was characterized primarily by cerebellar interactions with limbic and somatomotor systems. For each component, the number of significant edges within a given network pair was divided by the total number of significant edges in that component, yielding the relative proportion of edges assigned to each network pair.

The increased-connectivity component comprised 112 edges connecting 97 nodes (P_FWE_ < 0.05) and was primarily characterized by enhanced cortico-cerebellar connectivity, particularly involving limbic and somatomotor regions (Fig.2b), with additional but smaller contributions from other network pairs.

Expression of the increased and decreased components was strongly inversely related across individuals (r = -x0.78, P < 0.001).

### Cross-state consistency of the identified connectivity alterations

We next projected the two NBS-derived edge masks onto the original Fisher z-transformed matrices. Because these masks were defined in the discovery analysis, this analysis assesses cross-state consistency rather than providing an independent validation. For each participant and state, component-expression scores were calculated as the mean connectivity across the predefined edges of each component.

The increased cerebellar component showed higher mean connectivity in the psychiatric group in all seven states (all FDR-corrected P < 0.001; Fig. 3 and Table S4). The decreased limbic-somatomotor-salience component showed the opposite pattern, with lower connectivity in the psychiatric group in every state (all FDR-corrected P < 0.001).

**Fig. 3.**
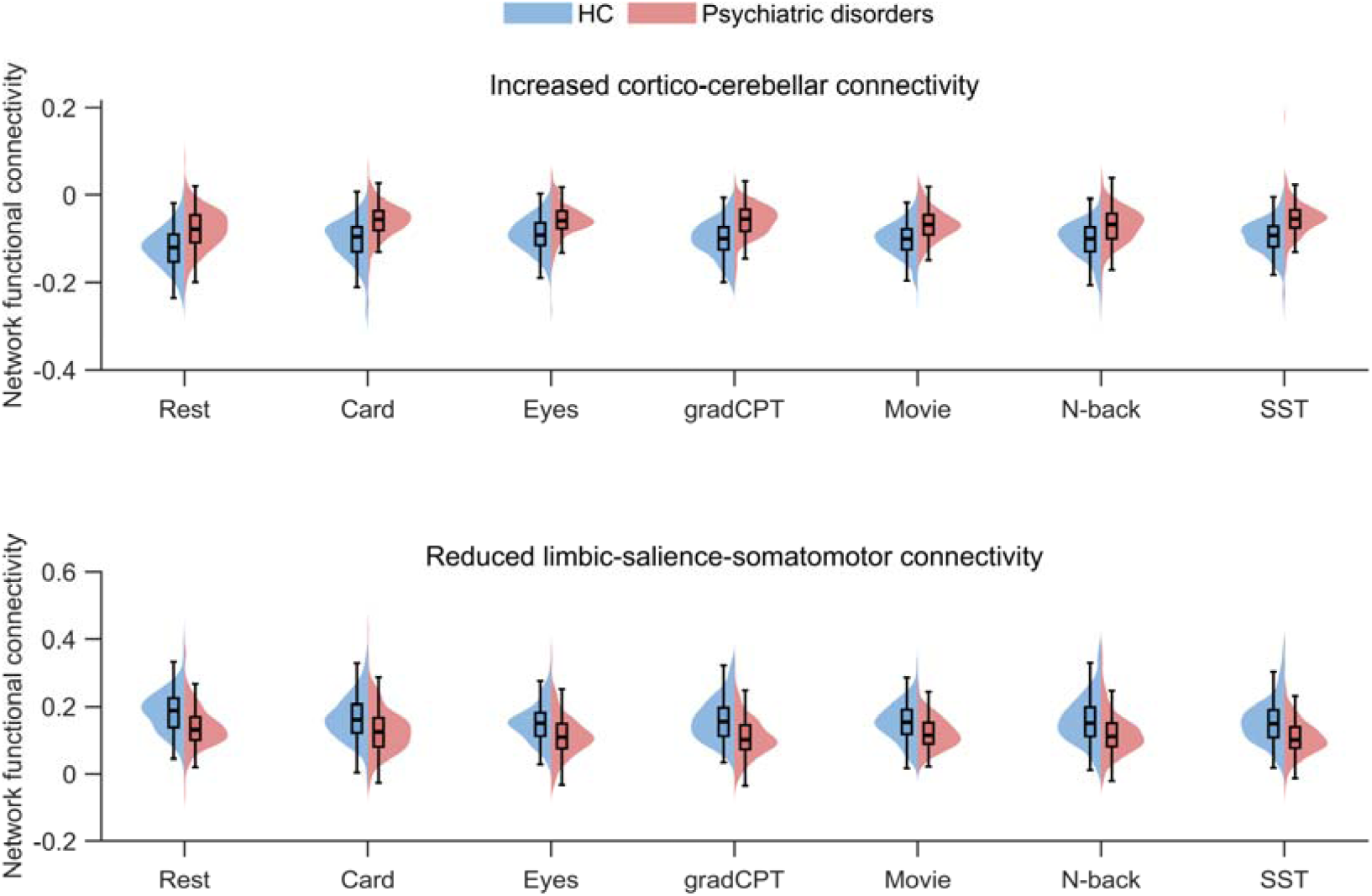
Cross-state consistency of the two connectivity components in the YaleNeuroConnect discovery cohort. Split violin plots show mean connectivity within the increased cortico-cerebellar component and the reduced limbic-salience-somatomotor component across one resting-state and six task conditions. The left and right halves represent healthy controls and participants with psychiatric disorders, respectively. Embedded boxplots indicate the median and interquartile range, with whiskers extending to the most extreme observations within 1.5 × the interquartile range. Component expression was calculated by averaging Fisher *z*-transformed connectivity across the predefined NBS edges within each component. Similar group differences were observed across all seven states. Functional states included resting state (Rest), Card Guessing (Card), Reading the Mind in the Eyes (Eyes), gradual-onset continuous performance task (gradCPT), movie watching (Movie), n-back working-memory task (N-back), and stop-signal task (SST).

The preservation of effect direction across cognitive contexts indicates that these components reflect cross-state features of functional organization rather than effects confined to a single task.

### Sensitivity analyses

The principal connectivity alterations in the YaleNeuroConnect cohort were robust across analyses restricted to White participants, non-smokers, medication-free participants, and individuals with a single psychiatric diagnosis, with patients consistently showing higher expression of the increased cortico-cerebellar component and lower expression of the reduced limbic–salience–somatomotor component. These findings support the robustness of the observed effects across demographic, smoking, medication, and diagnostic-comorbidity sensitivity analyses (Supplementary Table S5).

### Independent validation of the state-independent connectivity components

Next, generalizability of the state-independent connectivity alterations was evaluated independently in the UCLA cohort. PC1 explained 68.84% of the variance across seven functional states. The proportion of variance explained did not differ between the psychiatric and healthy-control groups after adjustment for age and sex (t = −0.782, P = 0.435). Using the Yale-derived edge masks, the psychiatric group showed higher expression of the increased cerebellar component (t = 4.063, P < 0.001, Fig. 4a) and lower expression of the limbic-somatomotor-salience component (t = −3.235, P = 0.0015; Fig. 4b). The same directional differences were also observed across individual UCLA functional states (Fig. 4c and Table S6).

**Fig. 4.**
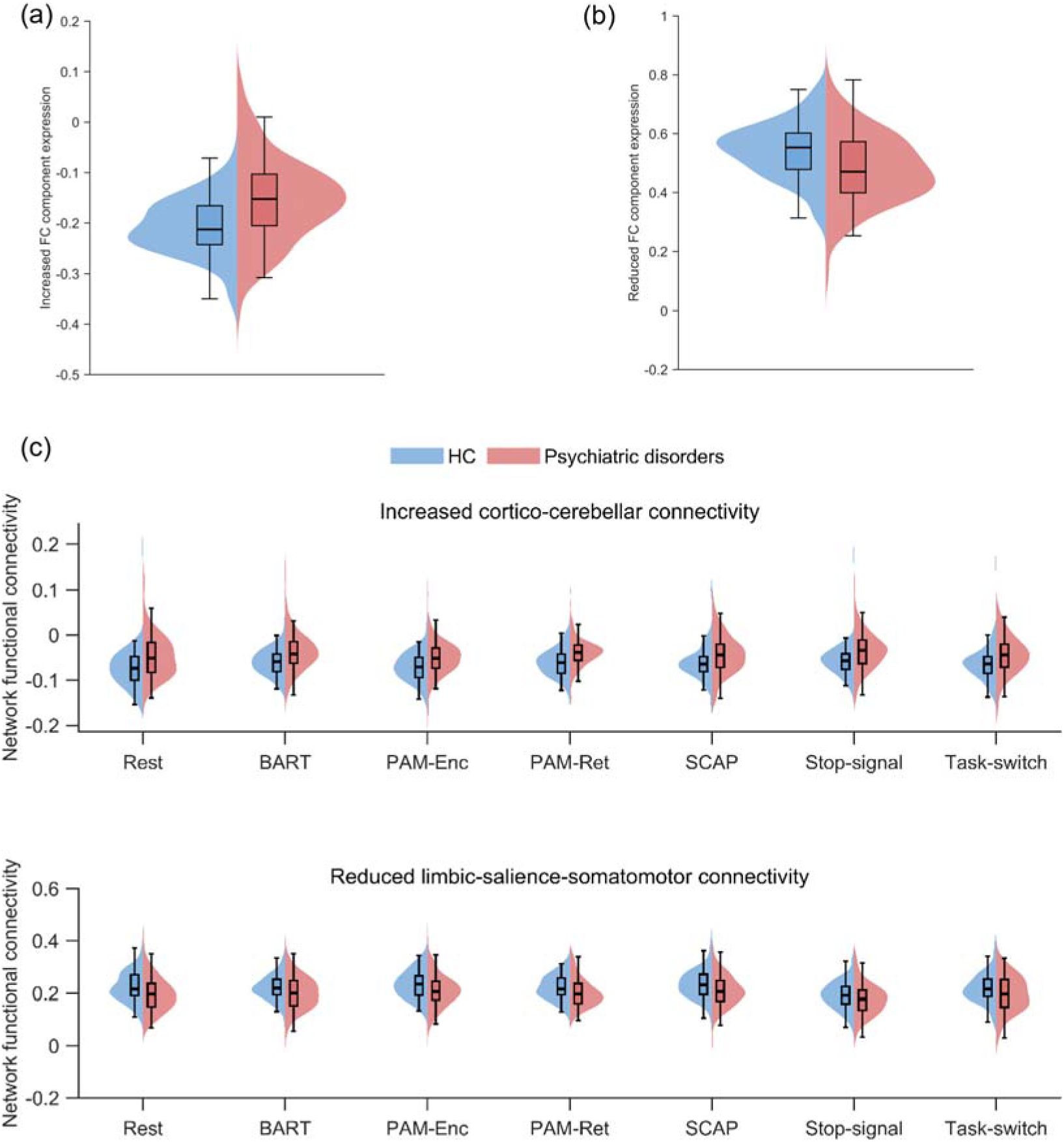
Validation of connectivity component alterations in the independent UCLA cohort. **a,b**, Participant-level expression of the increased cortico-cerebellar component (**a**) and reduced limbic–salience–somatomotor component (**b**) in healthy controls and individuals with psychiatric disorders. Group differences were tested using linear models adjusting for age, sex and education. **c**, Cross-state expression of the same components across one resting-state and six task conditions, showing consistent directional group differences across all seven states. Violin shapes show data distributions; embedded boxes indicate the interquartile range, horizontal lines the median, and whiskers the most extreme observations within 1.5 × the interquartile range. Functional states included resting state (Rest), Balloon Analogue Risk Task (BART), Paired Associative Memory encoding (PAM-Enc), Paired Associative Memory retrieval (PAM-Ret), Spatial Working Memory Capacity task (SCAP), Stop-Signal task, and Task-Switching task.

### Clinical relevance of the state-independent connectivity components

Within the psychiatric group in the Yale discovery cohort, the increased cerebellar component was associated with multiple clinical domains after FDR correction (Fig. 5 and Table S7). Greater component expression was associated with lower positive affect and with greater psychopathology and everyday executive difficulties. By contrast, the decreased limbic-somatomotor-salience component showed no significant associations with the available clinical or cognitive measures after FDR correction.

**Fig. 5.**
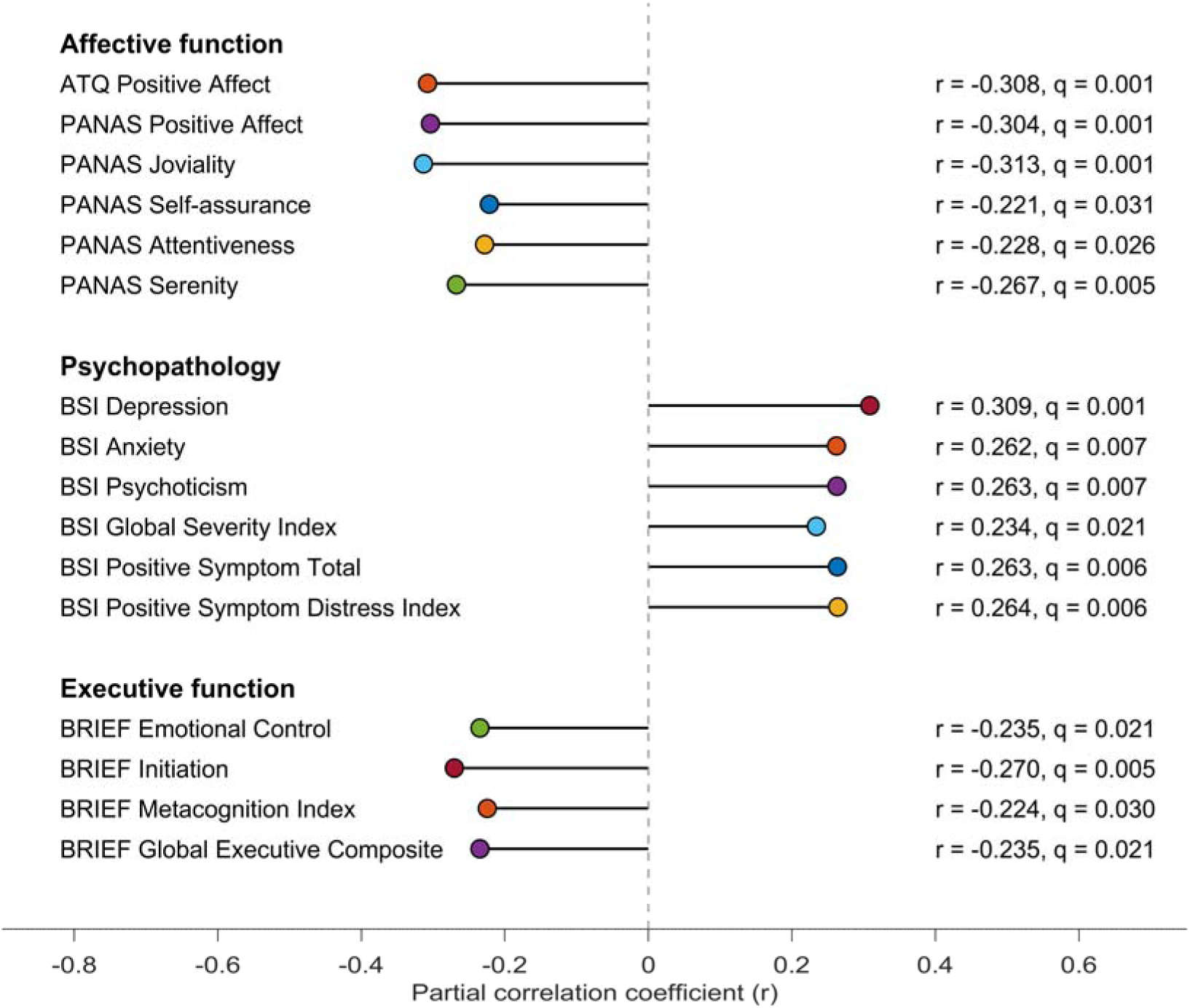
Dimensional clinical associations of the increased cerebellar connectivity component in the Yale psychiatric group. Partial correlations controlled for age and sex, and only associations surviving false-discovery-rate correction are shown. Greater component expression was related to lower positive affect, including lower joviality, serenity, attentiveness and broader positive-affect scores. It was also related to higher depression, anxiety and psychoticism scores, as well as greater global symptom severity and distress. Greater expression of the same component was additionally associated with more everyday executive difficulties, including poorer emotional control, initiation, metacognitive regulation and global executive functioning. BRIEF correlation coefficients were sign-reversed for visualization so that poorer executive functioning is displayed in the same direction as greater symptom burden.

The two components therefore differed not only in anatomical organization and direction of case-control alteration, but also in their relation to clinical variation. In the discovery cohort, stronger cerebellar coupling with limbic and somatomotor systems indexed a clinically varying dimension of affective, psychopathological and executive dysfunction, whereas the reduced limbic-somatomotor-salience component was expressed at the group level without detectable associations with the measured symptom dimensions.

## Discussion

There is growing recognition that psychiatric disorders share substantial neurobiological features despite considerable heterogeneity in their clinical manifestations. However, most functional neuroimaging studies have characterized these abnormalities under a single brain state, leaving it unclear which alterations represent stable features of psychiatric brain organization and how such features relate to individual clinical variation. By integrating functional connectivity across resting state and six task conditions, we identified two reproducible patterns of state-independent connectivity alteration in a diagnostically heterogeneous psychiatric population. One pattern was characterized by reduced connectivity among limbic, somatomotor and salience systems, whereas the other was dominated by increased cerebello-cortical connectivity, particularly along cerebellar-limbic and cerebellar-somatomotor connections. Both patterns were consistently expressed across functional states and generalized to an independent UCLA cohort. Importantly, only the cerebellar pattern was significant associated with affective, psychopathological and everyday executive difficulties. Together, these findings suggest that psychiatric disorders are associated with a stable reorganization of large-scale functional networks, within which different connectivity features may relate differently to common psychiatric abnormalities and individual clinical expression.

The reduced connectivity pattern mainly involved interactions among limbic, somatomotor and salience systems. These systems support affective processing, representation of bodily states and the detection and selection of behaviorally relevant information (35,36). The salience network, in particular, plays an important role in identifying biologically and behaviorally relevant internal and external events and coordinating responses across other large-scale systems (37,38). The limbic and somatomotor systems provide complementary representations of affective value, bodily states and action-related information. Reduced coordination among these systems may therefore indicate impaired integration of affective, interoceptive and sensorimotor information. Consistent with this interpretation, abnormalities involving salience, limbic and somatomotor networks have been reported across schizophrenia, bipolar disorder and other psychiatric conditions (6–8). The present findings extend this literature by showing that this pattern of dysconnectivity is not restricted to a single state, but remains evident across substantially different functional contexts.

The state-independent nature of this pattern may be particularly relevant for understanding psychiatric pathophysiology. Functional connectivity observed under a single condition reflects both relatively stable organization and transient responses to the current cognitive state. In contrast, an alteration that persists during reward processing, social cognition, sustained attention, naturalistic perception, working memory, inhibitory control and rest is less likely to be explained by abnormal engagement with one specific task (14). Reduced limbic-somatomotor-salience connectivity may therefore reflect a relatively stable disturbance in systems that coordinate internal states with ongoing environmental demands (39,40). Interestingly, this component did not show significant associations with the available affective, psychopathological or executive-function measures. This finding does not indicate that the component is clinically irrelevant. Rather, it raises the possibility that some neural abnormalities are expressed relatively broadly across psychiatric populations and may therefore be more closely related to common aspects of psychiatric dysfunction than to variation in current symptom severity (41–43).

A second pattern was characterized by stronger cerebellar coupling with limbic and somatomotor systems. The cerebellum has traditionally been associated with motor control, but increasing evidence indicates that it contributes to cognitive, affective and social processes through extensive interactions with cortical and subcortical systems (44,45). Cerebellar circuits are thought to support the coordination and adaptive adjustment of behavior by integrating ongoing internal and external information (46,47). In this context, enhanced cerebellar connectivity with limbic and somatomotor systems may reflect altered regulation of affective and bodily-state information in psychiatric disorders (47–49). Previous studies have reported cerebellar dysconnectivity in schizophrenia and bipolar disorder and demonstrated associations with clinical and cognitive abnormalities (9,50). State-independent cerebello-thalamo-cortical hyperconnectivity has also been linked to psychosis vulnerability and subsequent conversion (15). Our findings extend these observations to a diagnostically heterogeneous psychiatric population and indicate that abnormal cerebellar integration may represent a more general feature of psychiatric brain dysfunction.

Notably, the cerebellar pattern was also the component that showed consistent relationships with clinical variation. Stronger cerebellar coupling was associated with lower positive affect, higher depression, anxiety and psychoticism scores, greater global symptom burden, and more difficulties in everyday executive functioning (51–53). These associations extended across several clinical domains rather than corresponding to one particular diagnosis. This result is consistent with an emerging view that clinically meaningful neural variation may follow dimensional features of psychopathology that cut across conventional diagnostic categories (5,54). The cerebellar component may therefore capture a feature of functional brain organization that is particularly sensitive to variation in clinical expression (48,52,55). Importantly, the present findings do not establish whether stronger cerebellar connectivity represents a pathogenic process or a compensatory response to dysfunction elsewhere in the brain. Nevertheless, its association with multiple dimensions of clinical impairment suggests that cerebellar reorganization deserves greater attention in transdiagnostic models of psychiatric neurobiology.

The relationship between the two connectivity patterns is also informative. Their individual expression was strongly inversely correlated, suggesting that they should not be regarded simply as two independent abnormalities. Rather, reduced integration among limbic, somatomotor and salience systems occurred together with stronger cerebellar coupling to limbic and somatomotor systems (56,57). One possibility is that these changes reflect a coordinated redistribution of large-scale functional organization (58–60). Weaker integration among cortical systems involved in affective, salience-related and bodily processing may be accompanied by altered recruitment of cerebellar regulatory systems (56,61). Such reorganization could represent an attempt to maintain functional coordination in the presence of disrupted cortical network integration, although an alternative possibility is that excessive cerebellar coupling itself contributes to the persistence of maladaptive network states (47). The present findings suggest that psychiatric brain dysfunction may involve changes in the balance of interactions among large-scale systems rather than isolated increases or decreases in individual connections.

This distinction also has implications for understanding the relationship between psychiatric neurobiology and clinical heterogeneity. Neuroimaging studies frequently identify reproducible differences between psychiatric and healthy populations, yet these abnormalities often show only modest relationships with symptom severity or everyday functioning (62,63). The current results suggest that group-level abnormality and individual clinical variation may not be represented uniformly within the same altered connectome. Thus, some network features may reflect relatively broadly expressed psychiatric dysfunction, while others may provide more information about how that dysfunction is expressed in individual patients (64–66). These findings support a move from searching for a single transdiagnostic neural abnormality toward characterizing multiple interacting features of brain organization that contribute differently to psychiatric status and clinical variation.

Another important finding is the consistency of these connectivity patterns across functional states. Previous cross-paradigm studies have shown that connectivity shared across tasks and rest can capture stable individual characteristics and clinically relevant features of psychosis, including vulnerability, genetic risk and treatment response (12–15). Here, both psychiatric connectivity patterns showed the same direction of group difference across seven functional conditions in the discovery cohort and generalized to the UCLA cohort despite differences in diagnostic composition and task structure. These results suggest that the identified alterations are not strongly dependent on a particular cognitive context. The present study therefore extends cross-paradigm connectivity from previous disorder-focused applications to a broader transdiagnostic setting. In this context, state-independent connectivity may provide a useful way to identify functional abnormalities that are sufficiently stable to remain detectable as the brain moves between different cognitive states.

These findings may also have implications for biologically informed approaches to psychiatric classification. Current diagnostic systems remain essential for clinical practice, but considerable symptom overlap and within-diagnosis heterogeneity suggest that diagnostic categories alone provide an incomplete representation of underlying neurobiology (5,67,68). The present results do not suggest replacing conventional diagnosis with connectivity-based classification. Rather, state-independent functional organization may provide an additional level of characterization that complements diagnosis (15,61,69). In particular, the coexistence of broadly expressed network abnormalities and clinically varying connectivity features may provide a neurobiological basis for considering both common dysfunction and individual variation within psychiatric populations (64,65).

Strengths of this study include integrating multiple functional states, using a deeply phenotyped, diagnostically heterogeneous discovery cohort, and validating findings in an independent dataset with a different task battery and diagnostic composition. Several limitations remain. First, the state-independent architecture was derived from only one resting-state and six task conditions, and future studies incorporating broader cognitive and affective paradigms may yield a more comprehensive characterization of state-independent brain organization. Second, we focused on BOLD functional connectivity alone; future research should integrate multimodal imaging, electrophysiological, molecular and genetic measures to clarify the biological basis of the identified network alterations. Third, clinical analyses relied on the available affective, psychopathological and executive-function measures; more comprehensive phenotyping will better delineate the clinical relevance of these connectivity patterns. Fourth, both cohorts mainly comprised adults, and future studies across the lifespan are needed to establish developmental generalizability. Finally, this cross-sectional design cannot establish causality; longitudinal studies are required to determine whether these connectivity alterations precede, accompany or follow changes in psychiatric symptoms and functioning.

## Conclusion

In summary, we identified and independently validated two coordinated patterns of state-independent connectivity alteration in diagnostically heterogeneous psychiatric populations. Reduced connectivity among limbic, somatomotor and salience systems was consistently expressed across cognitive states, whereas increased cerebellar coupling with limbic and somatomotor systems additionally showed systematic associations with affective, psychopathological and everyday executive difficulties. These findings indicate that stable functional brain organization contains both broadly expressed psychiatric alterations and connectivity features more closely related to individual clinical variation, and that state-independent connectomics can help characterize how common neurobiological dysfunction and clinical heterogeneity coexist across psychiatric disorders.

## Acknowledgements

This work was supported by the National Natural Science Foundation of China (Grant Nos. 32300861 and 32671445), the Natural Science Foundation of Chongqing (CSTB2023NSCQ-MSX0896) and the Fundamental Research Funds for the Central Universities (SWU2509113).

## Data availability

The processed YaleNeuroConnect dataset and the UCLA Consortium for Neuropsychiatric Phenotyping (CNP) LA5c Study dataset are publicly available from OpenNeuro under accession numbers ds007286 and ds000030, respectively.

## Author Contribution

Yulin Wang and Debo Dong conceived and designed the study. Enhui Zhang, Anfu Deng, and Yanlong Hou performed the analyses. Bihua Xu and Shaolei Guo contributed to visualization of the results. Enhui Zhang, Ruoyun Zhang, Jiaxi Liao, and Yulin Wang interpreted the results. Enhui Zhang, Ruoyun Zhang, and Debo Dong wrote the original manuscript; Yulin Wang and Debo Dong supervised the study and obtained funding. All authors contributed to manuscript revision and approved the final version of the manuscript.

