## Supplementary materials for "State-independent functional connectomic architecture captures transdiagnostic network alterations and clinical variation"

**MRI preprocessing for YaleNeuroConnect cohort**

Structural and functional MRI data underwent standard preprocessing procedures. Brain extraction of the T1-weighted images was performed using optiBET, an optimized implementation based on the FSL framework. Functional images were motion-corrected in SPM12. Structural images were nonlinearly registered to MNI standard space using BioImage Suite, while functional-to-structural alignment was performed using tools from FSL and BioImage Suite. Subsequent preprocessing was conducted in BioImage Suite and included regression of mean white matter, cerebrospinal fluid, and grey matter signals, removal of linear, quadratic, and cubic trends, regression of 24 motion parameters, and temporal low-pass filtering using a Gaussian kernel (σ = 1.55). The preprocessing pipeline is publicly available at the YaleNeuroConnect Processing Pipeline repository (<https://github.com/YaleMRRC/YaleNeuroConnect_ProcessingPipeline>).

**MRI preprocessing for UCLA CNP cohort**

Anatomical and functional MRI data were preprocessed using fMRIPrep 24.1.1. Structural images underwent bias-field correction, brain extraction, tissue segmentation, and nonlinear normalization to MNI standard space. Functional images were corrected for head motion and co-registered to the corresponding structural images using boundary-based registration. Head motion parameters were estimated using FSL’s mcflirt, generating six rotation and translation parameters. The BOLD time series were further resampled into standard space, aligned with the MNI152NLin6Asym T1 template, using antsApplyTransforms with Lanczos interpolation to minimize smoothing effects. Nuisance regression was performed including the Friston 24-parameter head motion model, as well as mean signals from white matter, cerebrospinal fluid and global signal. The data were subsequently temporally low-pass filtered using a Gaussian kernel (σ = 1.55).

**MRI acquisition**

MRI data were acquired on a 3 T Siemens Prisma scanner with a 64-channel head coil. High-resolution T1-weighted structural images were collected using a three-dimensional MPRAGE sequence for anatomical registration (208 sagittal slices; repetition time = 2400 ms; echo time = 1.22 ms; flip angle = 8 degrees; slice thickness = 1 mm; in-plane resolution = 1 x 1 mm).

Functional images were acquired using a multiband gradient-echo echo-planar imaging sequence with whole-brain coverage (75 axial-oblique slices aligned parallel to the anterior commissure-posterior commissure plane). Acquisition parameters were repetition time = 1000 ms, echo time = 30 ms, flip angle = 55 degrees, slice thickness = 2 mm, multiband acceleration factor = 5 and in-plane resolution = 2 x 2 mm.

Table S1. Mental health diagnoses in the YaleNeuroConnect cohort, stratified by sex.

| **Diagnosis** | **Female** | **Male** | **Total** |
| --- | --- | --- | --- |
| All | 198 | 179 | 377 |
| No-mental-health diagnosis | 112 | 111 | 223 |
| Major depressive episode (MDE) | 53 | 30 | 83 |
| Generalized anxiety disorder (GAD) | 55 | 30 | 85 |
| Bipolar disorder | 18 | 22 | 40 |
| Alcohol use disorder (AUD) | 8 | 12 | 20 |
| Substance use disorder (SUD) | 7 | 18 | 25 |
| Panic episode | 27 | 14 | 41 |
| Post-traumatic stress disorder (PTSD) | 19 | 10 | 29 |
| Social anxiety disorder (SAD) | 7 | 5 | 12 |
| Obsessive compulsive disorder (OCD) | 12 | 9 | 21 |
| Autism spectrum disorder (ASD) | 1 | 1 | 2 |
| Attention-deficit/hyperactivity disorder (ADHD) | 19 | 26 | 45 |
| Agoraphobia | 10 | 5 | 15 |
| Psychotic episode/schizophrenia | 14 | 18 | 32 |
| Borderline personality disorder (BPD) | 8 | 1 | 9 |

Note: Diagnostic categories were not mutually exclusive; participants could meet criteria for more than one psychiatric disorder.

Table S2. Clinical and cognitive measures in YaleNeuroConnect dataset

| Assessment | Measure | Abbrevation | HC | Patient | T value | FDR q |
| --- | --- | --- | --- | --- | --- | --- |
| ATQ | Fear | atq_fear | 3.17 (0.89) | 3.98 (1.05) | 7.68308593 | 1.54386E-12 |
| ATQ | Frustration | atq_frustration | 3.30 (1.05) | 3.74 (1.05) | 3.887049485 | 0.000215057 |
| ATQ | Sadness | atq_sadness | 3.99 (0.95) | 4.69 (1.15) | 6.048105934 | 1.38901E-08 |
| ATQ | Discomfort | atq_discomfort | 3.69 (1.31) | 4.16 (1.30) | 3.361232985 | 0.001428475 |
| ATQ | Activation control | atq_activation | 5.04 (0.97) | 4.35 (1.08) | -6.259002773 | 4.21888E-09 |
| ATQ | Effortful attention | atq_attention | 4.52 (1.20) | 3.65 (1.23) | -6.679570227 | 4.12462E-10 |
| ATQ | Inhibitory control | atq_inhibitory | 4.68 (0.94) | 4.27 (0.96) | -4.03120302 | 0.000124009 |
| ATQ | Sociability | atq_sociability | 4.62 (1.22) | 4.23 (1.41) | -2.699221493 | 0.010706321 |
| ATQ | High intensity pleasure | atq_pleasure | 4.13 (1.08) | 4.13 (1.16) | 0.047631088 | 0.972852075 |
| ATQ | Positive affect | atq_positive | 4.68 (1.10) | 3.74 (1.26) | -7.30817824 | 1.34223E-11 |
| ATQ | Neutral perceptual sensitivity | atq_neutral | 4.72 (1.08) | 4.75 (1.11) | 0.249623304 | 0.840398505 |
| ATQ | Affective perceptual sensitivity | atq_affective | 4.52 (1.16) | 4.96 (1.06) | 3.730579101 | 0.000382262 |
| ATQ | Associative perceptual sensitivity | atq_associative | 4.32 (1.21) | 4.96 (1.21) | 4.957186569 | 2.59486E-06 |
| BRIEF | Inhibit | brief_inhibit | 11.00 (2.82) | 13.43 (3.55) | 7.061716735 | 5.59838E-11 |
| BRIEF | Shift | brief_shift | 8.40 (2.17) | 10.58 (2.79) | 8.157167193 | 7.95666E-14 |
| BRIEF | Emotional control | brief_emotional | 13.41 (3.98) | 16.81 (5.21) | 6.834531947 | 2.21816E-10 |
| BRIEF | Self-monitor | brief_self-monitor | 7.93 (2.28) | 9.05 (2.71) | 4.199120844 | 6.54975E-05 |
| BRIEF | Behavioral regulation index | brief_BRI | 40.75 (9.38) | 49.82 (11.71) | 7.991861923 | 2.08123E-13 |
| BRIEF | Initiate | brief_initiate | 11.45 (3.08) | 14.84 (3.91) | 8.996274317 | 5.6945E-16 |
| BRIEF | Working memory | brief_WM | 11.44 (3.22) | 14.75 (4.25) | 8.18973085 | 7.32892E-14 |
| BRIEF | Plan/organize | brief_plan/organize | 13.83 (3.61) | 16.73 (4.44) | 6.703919936 | 4.09752E-10 |
| BRIEF | Task monitor | brief_task | 8.88 (2.36) | 10.41 (2.58) | 5.80976562 | 3.94153E-08 |
| BRIEF | Organization of materials | brief_organization | 11.27 (3.04) | 13.66 (4.29) | 5.956755789 | 2.22247E-08 |
| BRIEF | Metacognition index | brief_MI | 56.84 (13.20) | 70.28 (16.69) | 8.311009776 | 3.36725E-14 |
| BRIEF | Global executive composite | brief_GEC | 97.62 (21.22) | 119.99 (25.67) | 8.862551173 | 9.9746E-16 |
| BSI | Somatization | bsi_somatization | 0.27 (0.92) | 0.68 (0.74) | 4.762855214 | 5.64257E-06 |
| BSI | Obsessive-compulsive | bsi_obsessive | 0.75 (0.71) | 1.76 (1.11) | 9.939162767 | 3.4542E-18 |
| BSI | Interpersonal sensitivity | bsi_interpersonal | 0.51 (0.70) | 1.35 (1.08) | 8.426497446 | 2.75892E-14 |
| BSI | Depression | bsi_depression | 0.48 (0.72) | 1.57 (1.28) | 9.557583309 | 5.76518E-17 |
| BSI | Anxiety | bsi_anxiety | 0.34 (0.48) | 1.11 (0.97) | 9.107082185 | 9.9746E-16 |
| BSI | Hostility | bsi_hostility | 0.33 (0.52) | 0.76 (0.90) | 5.303454847 | 6.48632E-07 |
| BSI | Phobic anxiety | bsi_phobic | 0.19 (0.40) | 0.66 (0.86) | 6.350075105 | 4.21888E-09 |
| BSI | Paranoid ideation | bsi_paranoid | 0.37 (0.55) | 0.86 (0.84) | 6.339301767 | 3.51151E-09 |
| BSI | Psychoticism | bsi_psychoticism | 0.30 (0.49) | 0.97 (0.87) | 8.610867823 | 1.41067E-14 |
| BSI | Global severity index | bsi_GSI | 0.39 (0.43) | 1.09 (0.75) | 10.47266989 | 1.97296E-19 |
| BSI | Positive symptom total | bsi_pst | 12.35 (10.10) | 25.36 (12.54) | 10.6733785 | 1.40629E-20 |
| BSI | Positive symptom distress index | bsi_psdi | 1.44 (2.35) | 1.91 (0.71) | 2.80240273 | 0.008145729 |
| IRI | Fantasy | iri_fantasy | 15.59 (5.55) | 16.61 (5.54) | 1.738313559 | 0.103878636 |
| IRI | Empathic concern | iri_empathic | 20.75 (4.39) | 20.96 (4.78) | 0.426881977 | 0.717610763 |
| IRI | Perspective taking | iri_pt | 19.68 (4.44) | 19.29 (4.56) | -0.805525502 | 0.467902284 |
| IRI | Personal distress | iri_pd | 9.51 (4.69) | 11.43 (5.24) | 3.616258918 | 0.000584891 |
| PANAS | Negative affect | panas_negative | 15.86 (6.10) | 21.78 (8.16) | 7.648101697 | 1.9684E-12 |
| PANAS | Fear | panas_fear | 9.13 (3.87) | 12.79 (5.94) | 6.721036497 | 4.45698E-10 |
| PANAS | Sadness | panas_sadness | 8.52 (4.07) | 12.29 (5.29) | 7.465658969 | 5.60959E-12 |
| PANAS | Guilt | panas_guilt | 8.76 (4.10) | 13.23 (6.96) | 7.168686368 | 4.79037E-11 |
| PANAS | Hostility | panas_hostility | 8.72 (3.25) | 11.07 (4.06) | 5.936147042 | 2.22247E-08 |
| PANAS | Shyness | panas_shyness | 5.94 (2.33) | 7.44 (3.31) | 4.854590488 | 4.52729E-06 |
| PANAS | Fatigue | panas_fatigue | 9.78 (3.51) | 12.44 (4.19) | 6.45678859 | 1.49625E-09 |
| PANAS | Positive affect | panas_positive | 33.77 (7.33) | 28.94 (8.75) | -5.605851992 | 1.18068E-07 |
| PANAS | Joviality | panas_joviality | 26.43 (6.48) | 21.23 (7.92) | -6.736620693 | 3.499E-10 |
| PANAS | Self-assurance | panas_self-assurance | 17.07 (5.04) | 14.36 (5.74) | -4.739924551 | 6.59241E-06 |
| PANAS | Attentiveness | panas_attentiveness | 13.96 (3.05) | 12.46 (3.43) | -4.344768521 | 3.5788E-05 |
| PANAS | Serenity | panas_serenity | 9.96 (2.74) | 8.51 (3.18) | -4.626175581 | 1.06662E-05 |
| PANAS | Surprise | panas_surprise | 6.12 (2.48) | 5.66 (2.36) | -1.816074612 | 0.090309752 |
| PSQI | Subjective sleep quality | psqi_quality | 0.93 (0.66) | 1.33 (0.77) | 5.248190055 | 6.80331E-07 |
| PSQI | Sleep latency | psqi_latency | 1.12 (0.88) | 1.71 (0.99) | 5.96070461 | 1.91102E-08 |
| PSQI | Sleep duration | psqi_duration | 0.65 (0.72) | 0.84 (0.92) | 2.189449014 | 0.040097835 |
| PSQI | Habitual sleep efficiency | psqi_efficiency | 0.37 (0.74) | 0.78 (1.04) | 4.183730837 | 7.15312E-05 |
| PSQI | Sleep disturbance | psqi_disturbance | 1.05 (0.53) | 1.38 (0.67) | 5.106015954 | 1.37701E-06 |
| PSQI | Use of sleep medication | psqi_medication | 0.35 (0.81) | 0.89 (1.20) | 4.811822679 | 5.44665E-06 |
| PSQI | Daytime dysfunction | psqi_daytime | 0.74 (0.73) | 1.25 (0.79) | 6.319351249 | 2.91912E-09 |
| PSQI | Global sleep quality score | psqi_global | 5.15 (3.00) | 8.25 (3.74) | 8.466200338 | 1.41067E-14 |
| PSS | Perceived stress scale | pss | 19.51 (8.03) | 27.61 (8.86) | 9.017614881 | 4.01964E-16 |
| BNT | Number correct | bnt_correct | 25.05 (4.39) | 25.97 (5.38) | 1.747088942 | 0.103592965 |
| BNT | Number of stimulus cues | bnt_stimulus cues | 2.31 (3.11) | 1.83 (3.34) | -1.415065744 | 0.1896732 |
| BNT | Number correct following stimulus cues | bnt_correct SC | 0.29 (0.63) | 0.26 (0.57) | -0.328428111 | 0.786505928 |
| BNT | Number of phoenomic cues | bnt_phoenomic cues | 8.22 (6.10) | 6.33 (5.04) | -3.248884258 | 0.002003209 |
| BNT | Number correct following phoenomic cues | bnt_correct PC | 3.22 (2.27) | 2.97 (2.22) | -1.016092773 | 0.349172326 |
| BNT | Total score | bnt_total | 51.81 (6.29) | 53.07 (6.35) | 1.847314137 | 0.085607655 |
| WRAML | Verbal learning | wraml_vl | 37.42 (9.76) | 33.89 (10.45) | -3.314295764 | 0.001649113 |
| WRAML | Verbal learning delayed recall | wraml_vl_cr_delay | 9.71 (3.44) | 8.66 (3.52) | -2.860269568 | 0.006994424 |
| WRAML | Verbal learning intrusion | wraml_vl_intrusion | 1.35 (1.94) | 1.25 (1.74) | -0.505952422 | 0.664926559 |
| WRAML | Finger windows | wraml_fw | 17.52 (3.54) | 16.58 (3.31) | -2.634477635 | 0.012387809 |
| WAIS | Symbol search | wais_symbol | 35.53 (9.86) | 32.59 (10.07) | -2.806836369 | 0.008099858 |
| WAIS | Coding | wais_coding | 74.61 (18.26) | 65.52 (18.35) | -4.728720966 | 6.59241E-06 |
| WAIS | Letter-number sequencing | wais_letter-number | 19.91 (3.14) | 19.49 (3.09) | -1.277852928 | 0.236328724 |
| WAIS | Cancellation | wais_cancellation | 37.94 (9.08) | 36.40 (8.86) | -1.641264035 | 0.125363222 |
| D-KEFS | Trail-making test: number sequencing | dkefs_TMT number | 28.99 (10.01) | 31.81 (11.05) | 2.531354851 | 0.016421955 |
| D-KEFS | Trail-making test: letter sequencing | dkefs_TMT letter | 27.48 (9.61) | 31.12 (14.83) | 2.682861332 | 0.011151201 |
| D-KEFS | Trail-making test: number-letter sequencing | dkefs_TMT number-letter | 70.75 (34.61) | 77.47 (42.61) | 1.622530325 | 0.128675222 |
| D-KEFS | Verbal fluency test: letter fluency | dkefs_VFT letter | 45.47 (12.61) | 43.87 (12.59) | -1.213258021 | 0.257354665 |
| D-KEFS | Verbal fluency test: category fluency | dkefs_VFT category | 45.15 (10.02) | 42.26 (10.08) | -2.724764507 | 0.010014146 |
| D-KEFS | Color-word interference test: color naming | dkefs_CW naming | 28.80 (6.37) | 29.70 (6.03) | 1.390003982 | 0.195911211 |
| D-KEFS | Color-word interference test: word reading | dkefs_CW reading | 21.36 (4.33) | 22.40 (4.97) | 2.102768301 | 0.048789112 |
| D-KEFS | Color-word interference test: inhibition | dkefs_CW inhibition | 48.69 (13.51) | 51.93 (16.05) | 2.050976216 | 0.054479764 |
| D-KEFS | Twenty-questions test: initial abstraction | dkefs_TQ initial | 35.32 (13.76) | 35.32 (14.27) | 0 | 1 |
| D-KEFS | Twenty-questions test: total questions asked | dkefs_TQ total | 26.27 (7.97) | 26.44 (6.89) | 0.216021416 | 0.847937863 |
| D-KEFS | Twenty-questions test: weighted achievement | dkefs_TQ weighted | 15.80 (2.65) | 15.74 (2.53) | -0.220321211 | 0.847937863 |
| WASI | Vocabulary | wasi_vocabulary | 40.53 (7.28) | 40.06 (6.71) | -0.637813764 | 0.575141064 |
| WASI | Matrix reasoning | wasi_matrix | 21.73 (4.27) | 21.18 (4.24) | -1.247661926 | 0.24581189 |

Abbreviations: ATQ, Adult Temperament Questionnaire; BRIEF-A, Behavior Rating Inventory of Executive Function–Adult Version; BSI, Brief Symptom Inventory; IRI, Interpersonal Reactivity Index; PANAS-X, Positive and Negative Affect Schedule–Expanded Form; PSQI, Pittsburgh Sleep Quality Index; PSS, Perceived Stress Scale; BNT, Boston Naming Test; WRAML, Wide Range Assessment of Memory and Learning; WAIS, Wechsler Adult Intelligence Scale; D-KEFS, Delis–Kaplan Executive Function System; WASI, Wechsler Abbreviated Scale of Intelligence.

Table S3. Demographic characteristics of the UCLA validation cohort.

| Variable | HC (n=74) | Patients(n=103) | Statistic | P value |
| --- | --- | --- | --- | --- |
| Age (years), mean ± SD | 29.69 ± 7.75 | 33.59 ± 9.38 | t=-3.025 | 0.0029 |
| Sex, Female n (%) | 41 (55.4%) | 63 (61.2%) | χ²=0.012 | 0.9126 |
| Race, White n (%) | 56 (75.7%) | 76 (73.8%) | χ²=0.012 | 0.9126 |
| School years, mean ± SD | 15.14 ± 1.51 | 13.94 ± 2.06 | t=4.444 | <0.0001 |

Table S4. Descriptive values and group-comparison statistics for the two connectivity components across functional states in YaleNeuroConnect cohort

| **Task** | **HC_mean** | **HC_SD** | **Patient_mean** | **Patient_SD** | **t_value** | **P_value** |
| --- | --- | --- | --- | --- | --- | --- |
| Increased FC | | | | | | |
| Rest | -0.1211 | 0.0466 | -0.0782 | 0.0460 | 8.261 | <0.001 |
| Card | -0.1043 | 0.0480 | -0.0599 | 0.0417 | 8.973 | <0.001 |
| Eyes | -0.0916 | 0.0392 | -0.0592 | 0.0403 | 7.467 | <0.001 |
| gradCPT | -0.1022 | 0.0420 | -0.0598 | 0.0366 | 9.377 | <0.001 |
| Movie | -0.1023 | 0.0383 | -0.0675 | 0.0350 | 8.182 | <0.001 |
| N-back | -0.1046 | 0.0417 | -0.0708 | 0.0411 | 7.243 | <0.001 |
| SST | -0.0976 | 0.0407 | -0.0560 | 0.0434 | 8.994 | <0.001 |
| Decreased FC | | | | | | |
| Rest | 0.1839 | 0.0616 | 0.1371 | 0.0613 | -7.215 | <0.001 |
| Card | 0.1654 | 0.0638 | 0.1302 | 0.0683 | -4.676 | <0.001 |
| Eyes | 0.1501 | 0.0550 | 0.1127 | 0.0633 | -5.933 | <0.001 |
| gradCPT | 0.1592 | 0.0618 | 0.1078 | 0.0591 | -7.230 | <0.001 |
| Movie | 0.1522 | 0.0557 | 0.1201 | 0.0451 | -5.332 | <0.001 |
| N-back | 0.1594 | 0.0693 | 0.1202 | 0.0651 | -4.936 | <0.001 |
| SST | 0.1536 | 0.0642 | 0.1091 | 0.0589 | -6.042 | <0.001 |

Notes: Functional states included resting state (Rest), Card Guessing (Card), Reading the Mind in the Eyes (Eyes), gradual-onset continuous performance task (gradCPT), movie watching (Movie), n-back working-memory task (N-back), and stop-signal task (SST).

Table S5. Sensitivity analyses of the principal connectivity component alterations in restricted subsamples

| **Task** | **HC_mean** | **HC_SD** | **Patient_mean** | **Patient_SD** | **t_value** | **P_value** |
| --- | --- | --- | --- | --- | --- | --- |
| **Only white** **(HC, n=103; Patient, n=111)** | | | | | | |
| Increased FC | -0.2621 | 0.0845 | -0.1659 | 0.0830 | -8.395 | <0.001 |
| Decreased FC | 0.4039 | 0.1046 | 0.3201 | 0.1149 | 5.587 | <0.001 |
| **Medication free (HC, n=158; Patient, n=30)** | | | | | | |
| Increased FC | -0.2829 | 0.0863 | -0.2070 | 0.0847 | -4.480 | <0.001 |
| Decreased FC | 0.4358 | 0.1160 | 0.3512 | 0.1595 | 2.769 | 0.009 |
| **Nonsmokers (HC, n=201; Patient, n=105)** | | | | | | |
| Increased FC | -0.2738 | 0.0808 | -0.1700 | 0.0806 | -11.293 | <0.001 |
| Decreased FC | 0.4289 | 0.1107 | 0.3145 | 0.1091 | 9.149 | <0.001 |
| **Single psychiatric diagnosis (HC, n=223; Patient, n=61)** | | | | | | |
| Increased FC | -0.2727 | 0.0825 | -0.2376 | 0.0976 | 2.894 | <0.005 |
| Decreased FC | 0.4225 | 0.1111 | 0.3855 | 0.1377 | −2.301 | 0.0223 |

Table S6. Descriptive values and group-comparison statistics for the two connectivity components across functional states in UCLA cohort

| Task | HC_mean | HC_SD | Patient_mean | Patient_SD | t_value | P_value |
| --- | --- | --- | --- | --- | --- | --- |
| **increased fc** |  |  |  |  |  |  |
| increased fc | -0.20688 | 0.053957 | -0.15473 | 0.082976 | 4.06261 | 7.35E-05 |
| rest | -0.07224 | 0.046307 | -0.04641 | 0.047228 | 3.093885 | 0.002304 |
| bart | -0.05964 | 0.028257 | -0.03669 | 0.042254 | 3.540965 | 0.000512 |
| pamenc | -0.07043 | 0.030347 | -0.05024 | 0.037594 | 3.394786 | 0.000852 |
| pamret | -0.06041 | 0.027711 | -0.03827 | 0.03563 | 3.95608 | 0.000111 |
| scap | -0.06151 | 0.035548 | -0.04278 | 0.044293 | 2.362374 | 0.01927 |
| stopsignal | -0.05576 | 0.038148 | -0.03486 | 0.042577 | 2.832901 | 0.005161 |
| taskswitch | -0.06263 | 0.040757 | -0.04486 | 0.043322 | 2.059362 | 0.040958 |
| **decreased fc** | | | | | | |
| decreased fc | 0.544568 | 0.093439 | 0.479809 | 0.116687 | -3.23527 | 0.001456 |
| rest | 0.223745 | 0.054381 | 0.194899 | 0.064687 | -2.9284 | 0.003866 |
| bart | 0.219677 | 0.046089 | 0.201179 | 0.058371 | -1.80882 | 0.072215 |
| pamenc | 0.232963 | 0.05133 | 0.209004 | 0.061271 | -2.24841 | 0.025812 |
| pamret | 0.221853 | 0.045195 | 0.202776 | 0.05428 | -1.86034 | 0.064535 |
| scap | 0.234305 | 0.050673 | 0.208435 | 0.059871 | -2.62506 | 0.009439 |
| stopsignal | 0.192781 | 0.051114 | 0.175686 | 0.060104 | -1.33098 | 0.184947 |
| taskswitch | 0.217825 | 0.056302 | 0.197461 | 0.065979 | -1.41444 | 0.159029 |

Notes: Functional states included resting state (Rest), Balloon Analogue Risk Task (BART), Paired Associative Memory encoding (PAM-Enc), Paired Associative Memory retrieval (PAM-Ret), Spatial Working Memory Capacity task (SCAP), Stop-Signal task, and Task-Switching task.

Table S7. Clinical associations of the increased cerebellar connectivity component within the psychiatric group in the Yale discovery cohort

| Network | Measure | r | p | p_fdr |
| --- | --- | --- | --- | --- |
| Increased FC | atq_fear | 0.177007518 | 0.034442407 | 0.129791652 |
| Increased FC | atq_frustration | 0.062126973 | 0.461047799 | 0.704540939 |
| Increased FC | atq_sadness | 0.192090637 | 0.021541509 | 0.088797061 |
| Increased FC | atq_discomf | 0.124122375 | 0.139681399 | 0.329362089 |
| Increased FC | atq_activation | -0.13292522 | 0.113504695 | 0.286413717 |
| Increased FC | atq_attn | -0.155564715 | 0.063557986 | 0.196121785 |
| Increased FC | atq_inhib | 0.113520872 | 0.177026871 | 0.389889746 |
| Increased FC | atq_sociab | -0.042170179 | 0.617019695 | 0.805717524 |
| Increased FC | atq_pleas | -0.030287447 | 0.719528638 | 0.865474204 |
| Increased FC | atq_pos | -0.30767761 | 0.000185421 | 0.001335034 |
| Increased FC | atq_neupercept | -0.023567852 | 0.779941085 | 0.886739794 |
| Increased FC | atq_affpercept | 0.01495707 | 0.859272231 | 0.930415607 |
| Increased FC | atq_assocsens | 0.14779449 | 0.078146938 | 0.223276966 |
| Increased FC | bnt_cr_spon | 0.071465746 | 0.38642877 | 0.628528723 |
| Increased FC | bnt_stimCues | 0.106938327 | 0.194259538 | 0.411299226 |
| Increased FC | bnt_cr_withStimCue | 0.174089041 | 0.044247836 | 0.149336446 |
| Increased FC | bnt_phonCue | -0.130557073 | 0.112505568 | 0.286413717 |
| Increased FC | bnt_cr_withPhonCue | -0.09730809 | 0.249301744 | 0.484255186 |
| Increased FC | bnt_totalScore | 0.053090459 | 0.523055093 | 0.750141131 |
| Increased FC | brief_inhib_raw | 0.098421233 | 0.227689131 | 0.455378263 |
| Increased FC | brief_shift_raw | 0.152051836 | 0.06147903 | 0.191899863 |
| Increased FC | brief_emo_raw | 0.234575642 | 0.003627388 | 0.020569273 |
| Increased FC | brief_sm_raw | -0.026868577 | 0.742470997 | 0.871227146 |
| Increased FC | brief_BRI_raw | 0.168343089 | 0.038157439 | 0.138288703 |
| Increased FC | brief_init_raw | 0.270277737 | 0.000758254 | 0.004761131 |
| Increased FC | brief_WM_raw | 0.192322485 | 0.017608887 | 0.076070393 |
| Increased FC | brief_plan_raw | 0.181453809 | 0.02526991 | 0.100336408 |
| Increased FC | brief_TM_raw | 0.168136804 | 0.039716454 | 0.141097929 |
| Increased FC | brief_org_raw | 0.069100039 | 0.397607641 | 0.636905166 |
| Increased FC | brief_MI_raw | 0.224431643 | 0.005761875 | 0.029917426 |
| Increased FC | brief_GEC_raw | 0.234620769 | 0.003853415 | 0.021233105 |
| Increased FC | bsi_som_raw | 0.085058027 | 0.299080062 | 0.548184471 |
| Increased FC | bsi_OC_raw | 0.16049452 | 0.04900115 | 0.160367398 |
| Increased FC | bsi_is_raw | 0.170172743 | 0.036707474 | 0.135767371 |
| Increased FC | bsi_dep_raw | 0.308772577 | 0.000114589 | 0.000883974 |
| Increased FC | bsi_anx_raw | 0.262170354 | 0.001191368 | 0.006917622 |
| Increased FC | bsi_hos_raw | 0.10607922 | 0.194854193 | 0.411299226 |
| Increased FC | bsi_phob_raw | 0.137751546 | 0.091660324 | 0.253828589 |
| Increased FC | bsi_par_raw | 0.084278349 | 0.30354758 | 0.551904692 |
| Increased FC | bsi_psy_raw | 0.2627655 | 0.001115858 | 0.006621576 |
| Increased FC | bsi_gsi_raw | 0.234181435 | 0.003802324 | 0.021167576 |
| Increased FC | bsi_pst_raw | 0.263451274 | 0.001081656 | 0.006489938 |
| Increased FC | bsi_psdi_raw | 0.264052504 | 0.001052463 | 0.006385734 |
| Increased FC | dkefs_tm_cond2_raw | 0.078133808 | 0.338663174 | 0.585579231 |
| Increased FC | dkefs_tm_cond3_raw | 0.042537751 | 0.602821745 | 0.801782616 |
| Increased FC | dkefs_tm_cond4_raw | -0.013205385 | 0.871723241 | 0.933483468 |
| Increased FC | dkefs_vf_cond1_raw | 0.019367346 | 0.812790384 | 0.914389182 |
| Increased FC | dkefs_vf_cond2_raw | -0.106014167 | 0.198164379 | 0.413161254 |
| Increased FC | dkefs_cw_cond1_raw | -0.122338379 | 0.133230469 | 0.317335265 |
| Increased FC | dkefs_cw_cond2_raw | -0.075628809 | 0.354422793 | 0.598088464 |
| Increased FC | dkefs_cw_cond3_raw | -0.068154697 | 0.404110921 | 0.639941047 |
| Increased FC | dkefs_20q_abstraction_raw | 0.085984488 | 0.292208532 | 0.542242637 |
| Increased FC | dkefs_20q_totalQ_raw | -0.15672245 | 0.053831949 | 0.171437824 |
| Increased FC | dkefs_20q_was_raw | 0.134396483 | 0.098785336 | 0.26742832 |
| Increased FC | iri_fs | 0.036834234 | 0.654511607 | 0.825785673 |
| Increased FC | iri_ec | -0.033192931 | 0.688791265 | 0.847260326 |
| Increased FC | iri_pt | 0.00646578 | 0.937196944 | 0.962141349 |
| Increased FC | iri_pd | 0.169427977 | 0.038859341 | 0.139127916 |
| Increased FC | panas_neg | 0.148398952 | 0.068061673 | 0.205325719 |
| Increased FC | panas_fear | 0.112515846 | 0.167548478 | 0.378561415 |
| Increased FC | panas_sad | 0.198926126 | 0.014014698 | 0.062544934 |
| Increased FC | panas_guilt | 0.162757868 | 0.04513133 | 0.150710867 |
| Increased FC | panas_hostility | 0.075988657 | 0.353746669 | 0.598088464 |
| Increased FC | panas_shyness | 0.047399515 | 0.563304718 | 0.7743072 |
| Increased FC | panas_fatigue | 0.16269668 | 0.04521326 | 0.150710867 |
| Increased FC | panas_pos | -0.303619261 | 0.000150772 | 0.001100228 |
| Increased FC | panas_jov | -0.313372726 | 8.46E-05 | 0.000661728 |
| Increased FC | panas_selfas | -0.221426421 | 0.006115768 | 0.030727253 |
| Increased FC | panas_attent | -0.228076111 | 0.004853829 | 0.025951163 |
| Increased FC | panas_serenity | -0.26740783 | 0.000866985 | 0.005320134 |
| Increased FC | panas_surprise | -0.109199649 | 0.180506062 | 0.394201633 |
| Increased FC | psqi_comp1_qual | 0.168261451 | 0.038904288 | 0.139127916 |
| Increased FC | psqi_comp2_lat | 0.148423271 | 0.068016023 | 0.205325719 |
| Increased FC | psqi_comp3_dur | -0.081514105 | 0.318114593 | 0.570703921 |
| Increased FC | psqi_comp4_eff | -0.090682796 | 0.271388712 | 0.51290532 |
| Increased FC | psqi_comp5_disturb | 0.10834527 | 0.183960762 | 0.394201633 |
| Increased FC | psqi_comp6_med | 0.202328714 | 0.012723367 | 0.057736285 |
| Increased FC | psqi_comp7_day | 0.134293042 | 0.099047526 | 0.26742832 |
| Increased FC | psqi_global | 0.136607128 | 0.097807505 | 0.26742832 |
| Increased FC | pss | 0.184017748 | 0.024184839 | 0.096739357 |
| Increased FC | wais_symbol_raw | -0.080241593 | 0.329010635 | 0.576265358 |
| Increased FC | wais_coding_raw | -0.078459792 | 0.338266786 | 0.585579231 |
| Increased FC | wais_LN_raw | 0.069492269 | 0.394927813 | 0.636600057 |
| Increased FC | wais_canc_tot_raw | -0.110936362 | 0.1736311 | 0.384265549 |
| Increased FC | wasi_vocab_raw | 0.082710959 | 0.31103707 | 0.559866725 |
| Increased FC | wasi_mr_raw | 0.029665948 | 0.71674905 | 0.865474204 |
| Increased FC | wraml_vl_cr_tot | -0.054285026 | 0.506539303 | 0.743351133 |
| Increased FC | wraml_vl_cr_delay | -0.077435112 | 0.343013403 | 0.588022976 |
| Increased FC | wraml_vl_intrusion | -0.16548251 | 0.042294833 | 0.145041919 |
| Increased FC | wraml_FW_cr | 0.036143409 | 0.658437176 | 0.82880204 |
| Decreased FC | atq_fear | -0.063304443 | 0.452579927 | 0.698266173 |
| Decreased FC | atq_frustration | -0.016429024 | 0.84558756 | 0.926468719 |
| Decreased FC | atq_sadness | -0.029143023 | 0.729707516 | 0.865474204 |
| Decreased FC | atq_discomf | -0.031728131 | 0.70678454 | 0.861543232 |
| Decreased FC | atq_activation | 0.0687275 | 0.4147214 | 0.654823263 |
| Decreased FC | atq_attn | 0.044682932 | 0.596177573 | 0.796871013 |
| Decreased FC | atq_inhib | -0.080463923 | 0.339419073 | 0.585579231 |
| Decreased FC | atq_sociab | -0.054209954 | 0.520197916 | 0.750141131 |
| Decreased FC | atq_pleas | -0.023651091 | 0.77918377 | 0.886739794 |
| Decreased FC | atq_pos | 0.13113807 | 0.11848371 | 0.293491759 |
| Decreased FC | atq_neupercept | 0.01008654 | 0.904830201 | 0.94834413 |
| Decreased FC | atq_affpercept | 0.004002252 | 0.962162461 | 0.973270972 |
| Decreased FC | atq_assocsens | -0.19567812 | 0.019174454 | 0.080892226 |
| Decreased FC | bnt_cr_spon | -0.035668968 | 0.665838887 | 0.830838152 |
| Decreased FC | bnt_stimCues | -0.140427518 | 0.087603854 | 0.246385841 |
| Decreased FC | bnt_cr_withStimCue | -0.181950335 | 0.035370458 | 0.131724463 |
| Decreased FC | bnt_phonCue | 0.027257568 | 0.741414904 | 0.871227146 |
| Decreased FC | bnt_cr_withPhonCue | -0.016874324 | 0.842014255 | 0.926044191 |
| Decreased FC | bnt_totalScore | -0.011731671 | 0.887846615 | 0.936400727 |
| Decreased FC | brief_inhib_raw | -0.000588224 | 0.994261467 | 0.996106108 |
| Decreased FC | brief_shift_raw | -0.045024948 | 0.581769468 | 0.790249645 |
| Decreased FC | brief_emo_raw | -0.116602187 | 0.152548929 | 0.350537964 |
| Decreased FC | brief_sm_raw | -0.021072985 | 0.796646826 | 0.901864331 |
| Decreased FC | brief_BRI_raw | -0.070517455 | 0.387974696 | 0.629148155 |
| Decreased FC | brief_init_raw | -0.147438281 | 0.069884994 | 0.20735108 |
| Decreased FC | brief_WM_raw | -0.078483177 | 0.336501137 | 0.585579231 |
| Decreased FC | brief_plan_raw | -0.166355975 | 0.040526788 | 0.142486188 |
| Decreased FC | brief_TM_raw | -0.129827597 | 0.113315629 | 0.286413717 |
| Decreased FC | brief_org_raw | -0.048886949 | 0.549772447 | 0.76773768 |
| Decreased FC | brief_MI_raw | -0.160561695 | 0.049672398 | 0.161584909 |
| Decreased FC | brief_GEC_raw | -0.152393492 | 0.062644387 | 0.194413615 |
| Decreased FC | bsi_som_raw | 0.04804301 | 0.558013292 | 0.772633789 |
| Decreased FC | bsi_OC_raw | -0.024608666 | 0.764231376 | 0.87805307 |
| Decreased FC | bsi_is_raw | -0.049447089 | 0.546551399 | 0.766591573 |
| Decreased FC | bsi_dep_raw | -0.102081286 | 0.212312098 | 0.429395255 |
| Decreased FC | bsi_anx_raw | -0.102871902 | 0.210313124 | 0.429131921 |
| Decreased FC | bsi_hos_raw | 0.011573251 | 0.887840378 | 0.936400727 |
| Decreased FC | bsi_phob_raw | -0.041187081 | 0.615581191 | 0.805717524 |
| Decreased FC | bsi_par_raw | 0.028019366 | 0.732716928 | 0.865474204 |
| Decreased FC | bsi_psy_raw | -0.132288705 | 0.105402733 | 0.276298427 |
| Decreased FC | bsi_gsi_raw | -0.053577669 | 0.513516517 | 0.747457159 |
| Decreased FC | bsi_pst_raw | -0.102412861 | 0.21082302 | 0.429131921 |
| Decreased FC | bsi_psdi_raw | -0.049879534 | 0.543044631 | 0.765650393 |
| Decreased FC | dkefs_tm_cond2_raw | -0.034717311 | 0.67111482 | 0.833108052 |
| Decreased FC | dkefs_tm_cond3_raw | -0.156632668 | 0.053971167 | 0.171437824 |
| Decreased FC | dkefs_tm_cond4_raw | -0.041416062 | 0.612425767 | 0.805717524 |
| Decreased FC | dkefs_vf_cond1_raw | -0.055410424 | 0.497754315 | 0.736403643 |
| Decreased FC | dkefs_vf_cond2_raw | 0.03927393 | 0.634398187 | 0.810673527 |
| Decreased FC | dkefs_cw_cond1_raw | 0.115883167 | 0.15511215 | 0.35490158 |
| Decreased FC | dkefs_cw_cond2_raw | 0.073070031 | 0.370986051 | 0.614516772 |
| Decreased FC | dkefs_cw_cond3_raw | 0.104622407 | 0.199580664 | 0.414513686 |
| Decreased FC | dkefs_20q_abstraction_raw | -0.097789385 | 0.230702568 | 0.459702535 |
| Decreased FC | dkefs_20q_totalQ_raw | 0.105589563 | 0.195433516 | 0.411299226 |
| Decreased FC | dkefs_20q_was_raw | -0.10531556 | 0.196602031 | 0.411492622 |
| Decreased FC | iri_fs | -0.080119109 | 0.329751844 | 0.576265358 |
| Decreased FC | iri_ec | 0.043514347 | 0.599489225 | 0.799318967 |
| Decreased FC | iri_pt | 0.007379261 | 0.928346352 | 0.956692805 |
| Decreased FC | iri_pd | -0.020725236 | 0.801906803 | 0.90591982 |
| Decreased FC | panas_neg | -0.024666225 | 0.76292562 | 0.87805307 |
| Decreased FC | panas_fear | -0.009133733 | 0.911076367 | 0.950845192 |
| Decreased FC | panas_sad | -0.123815594 | 0.12857282 | 0.314159832 |
| Decreased FC | panas_guilt | -0.053221492 | 0.514914932 | 0.747457159 |
| Decreased FC | panas_hostility | -0.002738835 | 0.973374937 | 0.978812786 |
| Decreased FC | panas_shyness | 0.041997385 | 0.608644001 | 0.805717524 |
| Decreased FC | panas_fatigue | -0.008571631 | 0.91652839 | 0.950845192 |
| Decreased FC | panas_pos | 0.194697806 | 0.016594232 | 0.072852726 |
| Decreased FC | panas_jov | 0.173048118 | 0.033008481 | 0.125525209 |
| Decreased FC | panas_selfas | 0.122285881 | 0.133398343 | 0.317335265 |
| Decreased FC | panas_attent | 0.123781273 | 0.129965077 | 0.315676982 |
| Decreased FC | panas_serenity | 0.14941276 | 0.06617969 | 0.203051321 |
| Decreased FC | panas_surprise | 0.093310537 | 0.252868592 | 0.485939643 |
| Decreased FC | psqi_comp1_qual | -0.0745424 | 0.363008729 | 0.604221606 |
| Decreased FC | psqi_comp2_lat | -0.086365397 | 0.290067941 | 0.541995462 |
| Decreased FC | psqi_comp3_dur | 0.144673575 | 0.075354704 | 0.217567017 |
| Decreased FC | psqi_comp4_eff | 0.056977268 | 0.490068529 | 0.727024741 |
| Decreased FC | psqi_comp5_disturb | -0.066438072 | 0.416079938 | 0.655052963 |
| Decreased FC | psqi_comp6_med | -0.12346549 | 0.130954979 | 0.315676982 |
| Decreased FC | psqi_comp7_day | -0.017200705 | 0.833410508 | 0.920330622 |
| Decreased FC | psqi_global | -0.050735056 | 0.540286505 | 0.763755793 |
| Decreased FC | pss | -0.037593892 | 0.647858595 | 0.823489716 |
| Decreased FC | wais_symbol_raw | 0.069776711 | 0.396174906 | 0.63670967 |
| Decreased FC | wais_coding_raw | 0.052289209 | 0.523709642 | 0.750141131 |
| Decreased FC | wais_LN_raw | 0.0117614 | 0.885648737 | 0.936400727 |
| Decreased FC | wais_canc_tot_raw | 0.145948536 | 0.072790797 | 0.212470434 |
| Decreased FC | wasi_vocab_raw | -0.028302044 | 0.729252556 | 0.865474204 |
| Decreased FC | wasi_mr_raw | 0.040243114 | 0.622539642 | 0.805717524 |
| Decreased FC | wraml_vl_cr_tot | -0.02990239 | 0.714588995 | 0.865474204 |
| Decreased FC | wraml_vl_cr_delay | 0.003849706 | 0.962456851 | 0.973270972 |
| Decreased FC | wraml_vl_intrusion | 0.094583226 | 0.248011928 | 0.483488958 |
| Decreased FC | wraml_FW_cr | 0.023002363 | 0.778489238 | 0.886739794 |

Abbreviations: ATQ, Adult Temperament Questionnaire; BRIEF-A, Behavior Rating Inventory of Executive Function–Adult Version; BSI, Brief Symptom Inventory; IRI, Interpersonal Reactivity Index; PANAS-X, Positive and Negative Affect Schedule–Expanded Form; PSQI, Pittsburgh Sleep Quality Index; PSS, Perceived Stress Scale; BNT, Boston Naming Test; WRAML, Wide Range Assessment of Memory and Learning; WAIS, Wechsler Adult Intelligence Scale; D-KEFS, Delis–Kaplan Executive Function System; WASI, Wechsler Abbreviated Scale of Intelligence.
